# Can social media forums serve as real-world data for nutraceuticals? Concordance between clinically supported and Reddit-reported ingredient benefits

**DOI:** 10.64898/2026.06.26.26356690

**Authors:** Anastasios Gkountakos, Christos Goulas, Yiannis Kourmpetis

**Affiliations:** Siftlink SA, Epalinges, Switzerland

**Keywords:** real-world evidence, real-world data, nutraceuticals, social media, Reddit, natural language processing, clinical trials, hypothesis generation

## Abstract

**Background:** Randomized controlled trials (RCTs) remain the reference standard for establishing clinical efficacy for candidate drugs, but their restrictive eligibility criteria limit generalizability, and for dietary supplements the body of well-powered RCTs is small relative to the number of proposed health claims. Self-reported real-world data (RWD) by patients in social media are increasingly recognized as a complementary evidence source, yet their value in the nutraceutical domain is largely uncharacterized.

**Objective:** To test whether benefits that users positively report for natural ingredients on Reddit are statistically associated with benefits demonstrated for those ingredients in the clinical-trial literature, and to characterize what a community corpus can reveal about application areas and side effects.

**Methods:** We assembled a cross-sectional corpus of 216,350 distinct comments from 86 supplement-related subreddits (January 2022 - April 2026), covering 329 canonical ingredients and a shared, MeSH/MedDRA-aligned vocabulary of 581 benefit terms. Comment-level directionality (benefit/no-effect/irrelevant) was assigned by a natural-language-inference model. The independence of clinical efficacy and Reddit endorsement was tested with 2×2 chi-squared and Fisher exact tests across a sweep of endorsement thresholds, plus per-ingredient tests with Bonferroni correction.

**Results:** Of 31,359 unique (ingredient, benefit) pairs, 2,508 were efficacy-demonstrated under the primary definition. The two signals were statistically associated. The association strengthened monotonically with endorsement volume, reaching OR 3.06 at ≥20 comments and 7.14 at ≥100. Among heavily-endorsed benefits (≥100 comments) 38% were RCT-demonstrated, versus ∼9% overall. Community attention concentrated on sleep, skin and anxiety, whereas the trial literature concentrated on cardiovascular, pain, metabolic and mood indications. The most discussed and clinically validated nutraceutical-health claim pairs include melatonin-sleep cycle and quality, ginger with gastrointestinal relief and zinc with skin conditions. The concordance analysis revealed that 81% (573/711) of clinically proved nutraceutical-health claim pairs were in agreement with the reported experience of the Reddit community.

**Conclusions:** Community endorsement and clinical efficacy are independent at the single-comment noise floor but positively concordant once a benefit is endorsed by multiple users, with concordance rising with consensus. Reddit-derived RWD is a useful hypothesis-generating prior for nutraceutical efficacy and a complementary, though not confirmatory, signal for prioritizing ingredients and indications for formal clinical study taking into consideration personalized characteristics.

## 1. Introduction

Randomized controlled trials (RCTs) are prospective studies which represent the highest standard for ensuring rigorous control, exploring causality and minimizing bias when assessing efficacy in the clinical setting [1]. However, RCTs are designed around strict eligibility criteria, so their conclusions are confined to specific settings and patient profiles and are generalized poorly to the broader real-world population. Moreover, RCTs are usually of too short duration for identifying longer-term adverse events, compliance to treatment or treatment discontinuation [2, 3].

Real-world data (RWD) refers to health and patient data originated from sources beyond RCTs or other controlled research settings. When RWD are systematically analyzed under the scope of scientific or clinical hypotheses might lead to novel clinical insights, the so-called real-world evidence (RWE) [4]. In recent years, RWD and RWE have gained increasing attention from health authorities and the pharmaceutical industry. As a result, RWD and RWE are increasingly being incorporated into regulatory decisions as a strategy for capturing extended effectiveness and safety evidence on approved products [5]. This long-term assessment capability can help bridge the gap between RCTs and routine clinical practice [6]. Interestingly, both the U.S. Food and Drug Administration (FDA) and the European Medicines Agency (EMA) have acknowledged the value of RWD/RWE and are developing frameworks for their appropriate handling [7]. Notably, Purpura et al. reported that 31% of FDA approvals between 2019 and 2021 incorporated RWE generated during routine clinical use [8].

RWE can be generated prospectively or retrospectively from observational studies and a variety of RWD sources [2], including electronic health records, insurance claims systems, wearable/mobile devices, and patient-led platforms. Owing to their high prevalence and real-time nature, social media platforms have emerged as a rich source of RWD, capturing authentic patient experience which could be translated into clinically relevant hypotheses [9]. Content from platforms such as Reddit and X, analyzed with AI tooling, has been used to detect potential drug-safety signals and generate hypotheses regarding previously misidentified indications or adverse effects; for example, for anticoagulants and GLP-1 receptor agonists [10, 11].

Nutraceuticals are naturally derived bioactive products, including dietary supplements, functional foods, probiotics and herbal products that provide health benefits beyond basic nutrition. They are widely used to support wellness and prevent disease, and their global market has grown rapidly across the food and pharmaceutical sectors [12]. Although numerous nutraceuticals are promoted with health-related claims, the evidence base supporting many of these claims remains limited [13]. Large, adequately powered, and independently replicated RCTs are generally uncommon, and much of the supporting information derives from preclinical work, small and underpowered studies or observational correlations [14, 15].

Existing RWE regarding both efficacy and adverse events of non-prescription products such as nutraceuticals has largely consisted of cross-sectional surveys spanning diverse indications and settings [16]. However, nutraceuticals are supported by significantly less RWD, predominantly because nutraceuticals are consumed extensively without physician prescription and timely documentation of RWD remains challenging due to limited access to healthcare databases, less nutraceutical-dedicated registries, misinformation regarding benefits and adverse events, etc [17]. Moreover, the efficacy of nutraceuticals is influenced by personalized factors, including genetic, metabolic and lifestyle characteristics.

In this study we explore the potential of social media-derived RWD and RWE for the nutraceutical domain, together with the statistical challenges affecting the integrity of data collection and analysis. We develop a framework that questions whether social-media-derived RWE is suitable for generating clinical hypotheses about supporting claims and unexpected effects across a wide population, and we position this stepwise approach as a complementary strategy to conventional experimental research. Collectively, we address three questions: (i) what do the two data sources describe observationally; (ii) are the clinically demonstrated benefits statistically associated and aligned with community-reported benefits; and (iii) which are the most discussed nutraceutical-health claim pairs and whether their associations are reflected on clinical trials.

## 2. Materials and methods

### 2.1 Study design and data sources

We conducted a cross-sectional study of self-reported personal experience with natural ingredients using Reddit data spanning January 2022 to April 2026. Three primary inputs were combined: a set of Reddit comments, ClinicalTrials and a set of Pubmed articles and a derived trial to benefit match table. Both the Reddit output and the clinical match table express effects on the same controlled benefit catalogue (e.g. “Improved Sleep Quality”, “Allergic Diseases (General)”), and this shared MeSH/MedDRA-aligned vocabulary is the join key that makes the association test possible.

### 2.2 Reddit corpus construction and directionality

For each comment we extracted the ingredient mentioned, the asserted effect (benefit), the normalized benefit term and a direction. Directionality was derived from the natural language relationship extraction (NLRE) based label of the hypothesis *“Taking {ingredient X} helped with {benefit Y}”*: support was coded as a positive report (it helped), contradiction as a no-effect/refutation report, and neutral as an irrelevant mention. A distinct “made it worse / harm” axis is not represented in this benefit-framed NLRE and is handled by a separate side-effects pipeline; within the benefits dataset the only negative-direction signal is the no-effect count, retained for inspection but not used to define a positive report. To avoid inflating support, we counted distinct comments asserting a benefit rather than raw mention rows.

### 2.3 Clinical evidence extraction

We analyzed the ClinicalTrials.gov and Pubmed corpora to identify clinical trials that use nutraceutical ingredients as intervention factors. Our analysis has yielded 13,875 distinct Clinical Trials, 12,859 scientific publications and 22,692 ingredient-benefit pairs.

### 2.4 Benefit-vocabulary harmonization

The 581 distinct benefit terms were grouped into 21 application areas by a local Biology optimized Large Language Model (LLM), so that, for example, insomnia, sleep-cycle regulation and improved sleep quality all map to “Sleep & insomnia” to NCBI MeSH, rebuilding a clean benefit to MeSH map.

### 2.5 Defining clinical support

Given that many of the clinical trials do not post results and even more posted or published results do not automatically imply statistically significant findings we used two filters to consider an ingredient-benefit pair as clinically supported:

1. Published results. At least one surviving trial with posted results in any direction (4,111 pairs).
2. Efficacy demonstrated i.e. at least one trial reporting a structured primary outcome with p-value < 0.05 (3,342 pairs).

### 2.6 Statistical analysis

For each ingredient-benefit pair we defined clinically supported (efficacy-demonstrated, primary definition) and Reddit found (distinct positive comments ≥ cutoff), and tabulated the 2×2 contingency table with cells A (both), B (clinical only), C (Reddit only) and D (neither). We report the Pearson χ² statistic (Yates-corrected), the Fisher exact test, the odds ratio and the φ effect size, at the primary cutoff and across a sweep of cutoffs. Because per-ingredient counts are expected to be small, the pooled all-pairs test is the primary analysis; per-ingredient tests are reported as an exploratory panel with Bonferroni correction (α = 0.05 / number of testable ingredients).

## 3. Results

The results are organized into three parts that mirror the study questions: (1) an observational description of the two data sources; (2) the potential statistically significant association between clinically proven benefits and self-reported consumer experience; and (3) the identification and the clinical association of the most discussed nutraceutical-health claim pairs.

### 3.1 Observational description of the data sources

The Reddit corpus comprised 216,350 distinct comments across 285 ingredients and 499 benefit terms; the clinical corpus comprised 102,726 trial records (median enrollment 61), of which 15,760 (∼15%) had posted results, with a phase mix dominated by observational/NA designs (65,521) followed by Phase 2 (13,217), Phase 1 (8,638), Phase 3 (7,694) and Phase 4 (6,301).

#### 3.1.1 Where community attention concentrates

Reddit users’ attention is heavily concentrated on sleep health/quality and physical appearance (skin, hair and anti-aging) with 65,672 and 35,707 distinct comments, respectively. Then, the next most frequently discussed topics concern anxiety & stress (13,674), nutrient deficiency (11,051) and digestion (7,601) (Figure 1).

**Figure 1.**
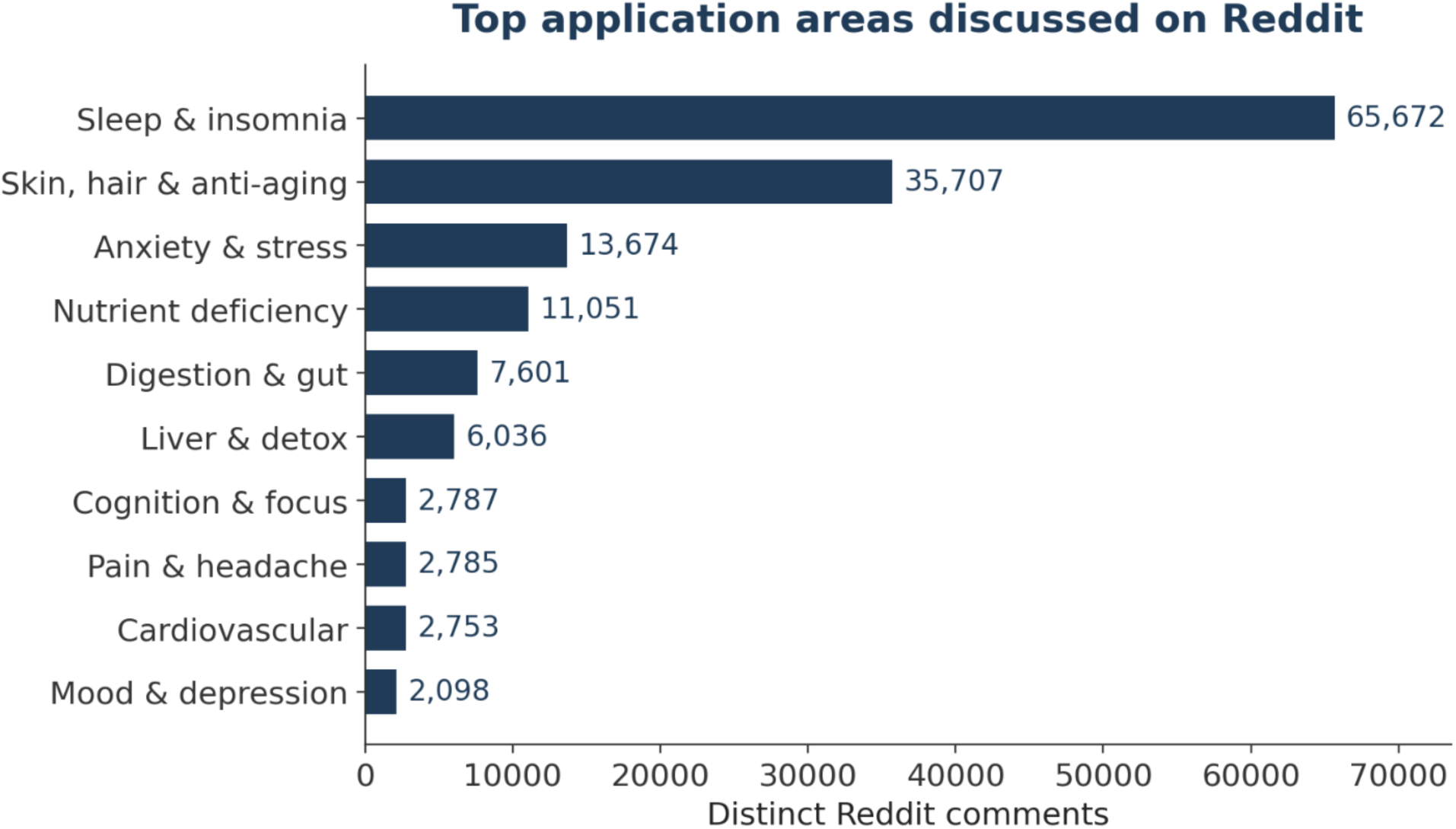
Distinct Reddit comments per application area (top ten).

The distribution of ingredient-containing reports is also highly heavy-tailed at the ingredient level: the two most-discussed ingredients, caffeine (67,684 distinct comments) and melatonin (56,243), together account for roughly 57% of all distinct comments (Figure 2). Application-area composition acts as a fingerprint of perceived use: melatonin is almost single-purpose (97% sleep), whereas zinc and L-theanine are multi-purpose (L-theanine split ∼43% anxiety / ∼42% sleep).

**Figure 2.**
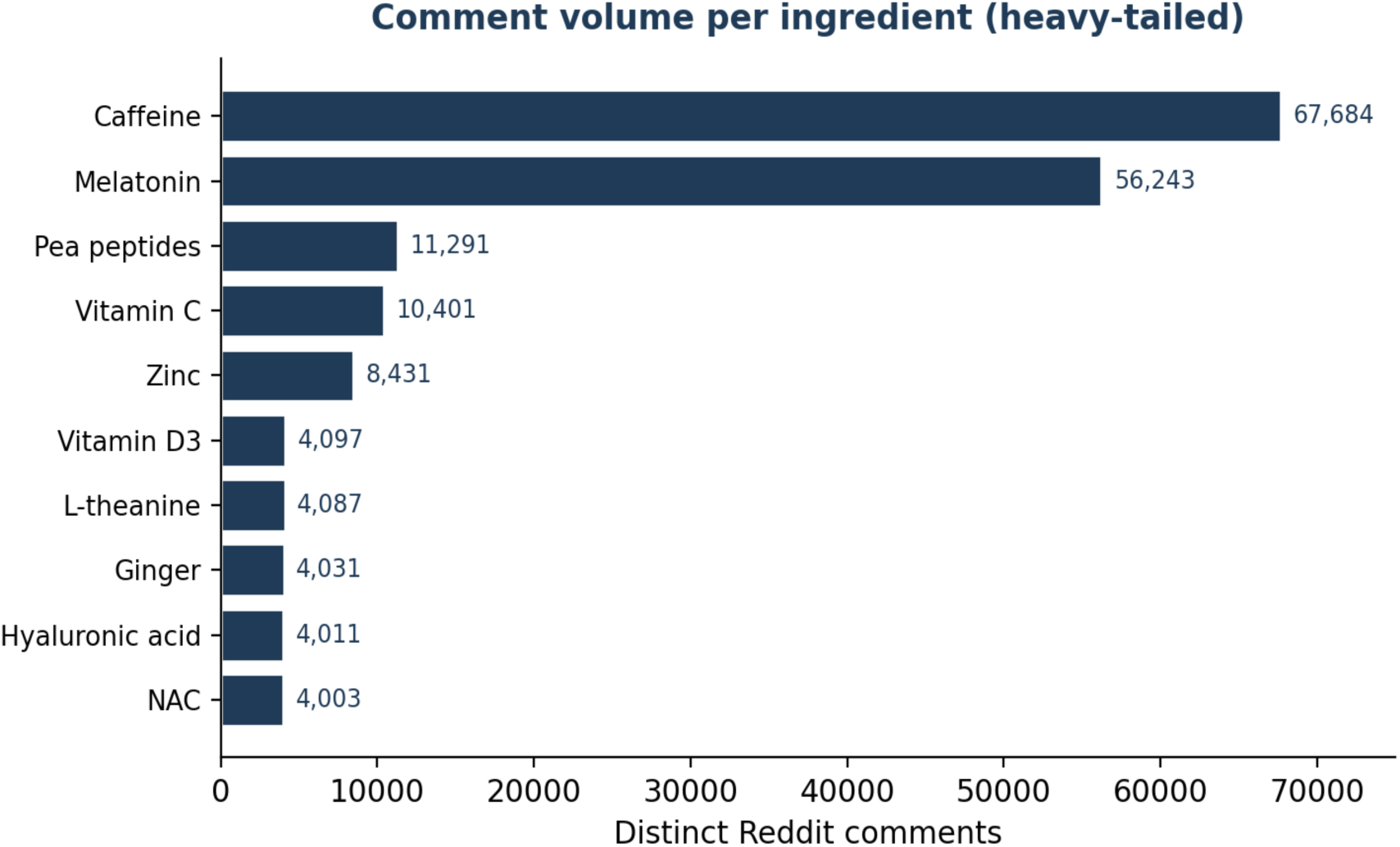
Distinct Reddit comments per ingredient (top ten).

#### 3.1.2 Discussion versus research: the attention gap

Mapping trials onto the same application areas reveals a sharp mismatch between what the community discusses and what is studied (Figure 3). The community over-indexes on sleep (39.2% of comments vs 3.0% of trials), skin (21.3% vs 4.8%) and anxiety (8.2% vs 5.9%), whereas the trial literature focuses on cardiovascular (1.6% vs 10.1%), pain (1.7% vs 6.3%), mood (1.5% vs 5.6%) and cognition. This gap demonstrates why low-volume Reddit pairs need not align with the clinical evidence base and it is precisely what the association analysis quantifies pair-by-pair.

**Figure 3.**
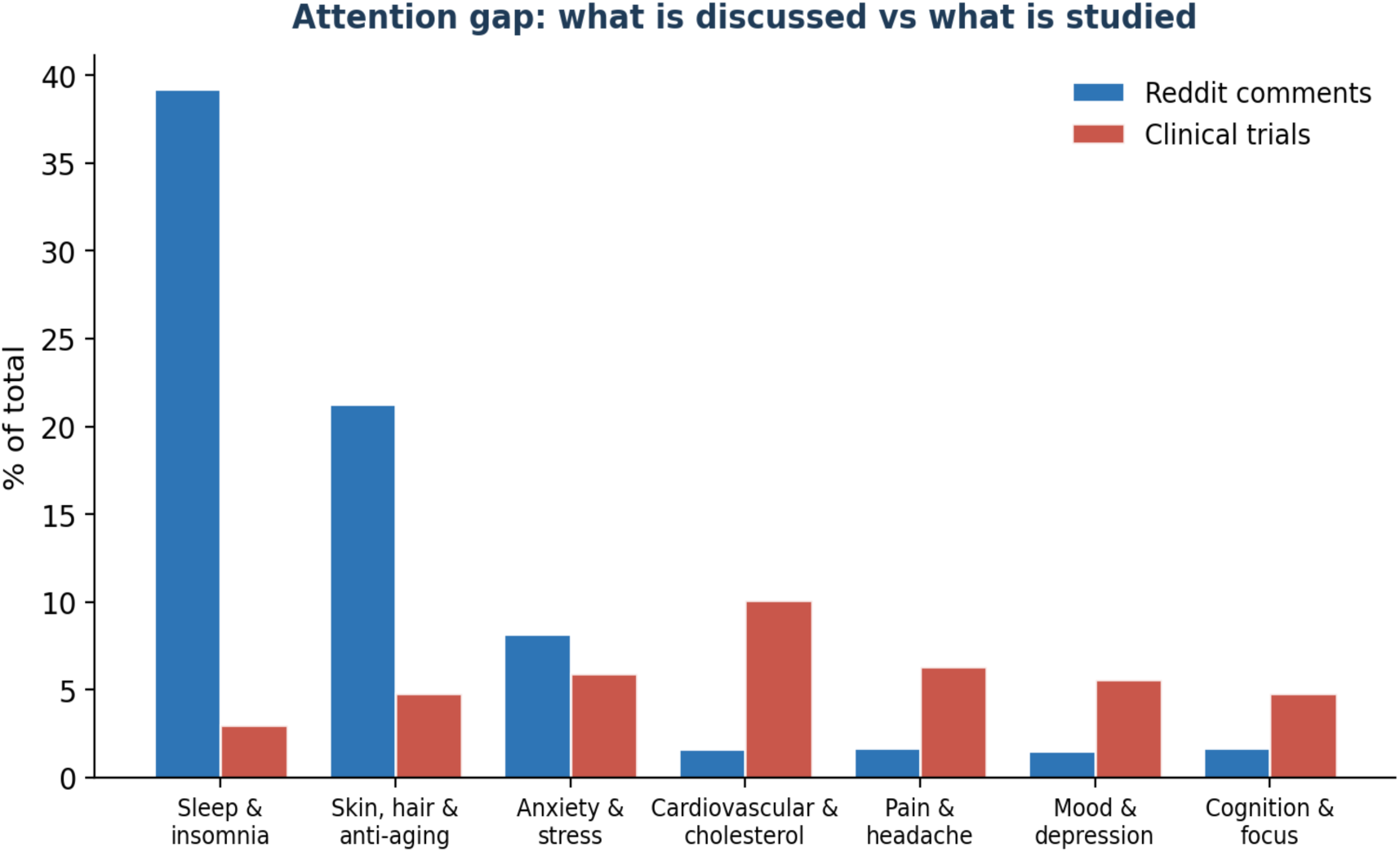
Share of Reddit comments versus share of clinical trials by application area. The community and the trial literature emphasize different indications.

### 3.2 Statistical association between clinical efficacy and community endorsement

Of the 31,359 unique (ingredient, benefit) pairs, 3,342 qualified as efficacy-demonstrated under the primary definition. At the primary Reddit threshold (≥1 positive comment) the 2×2 table was as shown in Table 1.

**Table 1.**
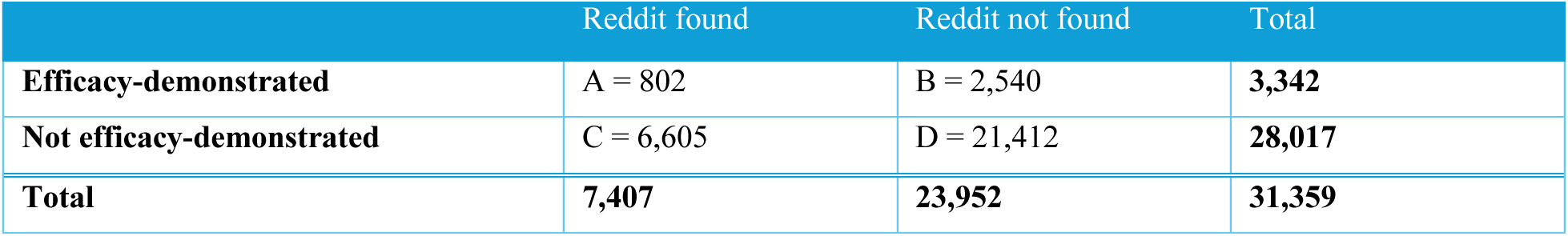
Primary 2×2 contingency table (efficacy-demonstrated, de-biased + spurious-cleaned; Reddit cutoff ≥1). Odds ratio 1.02; χ² p = 0.60; Fisher p = 0.59 - independence cannot be rejected at the single-comment noise floor.

|  | Reddit found | Reddit not found | Total |
| --- | --- | --- | --- |
| **Efficacy-demonstrated** | A = 802 | B = 2,540 | **3,342** |
| **Not efficacy-demonstrated** | C = 6,605 | D = 21,412 | **28,017** |
| **Total** | **7,407** | **23,952** | **31,359** |

At this lowest threshold the two signals are **statistically independent** (OR 1.02; χ² p = 0.60). A single passing endorsement is a weak, noisy signal—many benefits attract one comment regardless of clinical evidence—so the “found” set is dominated by noise. Interpreted as overlap rates, 24.0% of efficacy-demonstrated benefits are positively discussed on Reddit, and 10.8% of Reddit-endorsed benefits are backed by a trial demonstrating significant benefit for that exact indication.

#### 3.2.1 Endorsement-threshold sweep

Requiring more endorsing comments filters the noise and reveals a genuine, positive (concordant) association that is highly significant from cutoff ≥2 onward and strengthens monotonically with endorsement volume (Table 2).

**Table 2.** Cutoff sweep (primary clinical-support definition). The odds ratio is significantly positive at every threshold ≥1, rising to ∼9.8 at ≥300 comments. Cell counts shrink at high cutoffs, widening confidence intervals even though significance holds.

| Cutoff | A | B | C | D | Odds_ratio | Fisher_p | Direction |
| --- | --- | --- | --- | --- | --- | --- | --- |
| $\geq 1$ | 684 | 1824 | 6723 | 22128 | 1.23 | 1.0e-05 | positive |
| $\geq 2$ | 475 | 2033 | 3368 | 25483 | 1.77 | 1.1e-23 | positive |
| $\geq 3$ | 379 | 2129 | 2382 | 26469 | 1.98 | 7.4e-27 | positive |
| $\geq 4$ | 330 | 2178 | 1854 | 26997 | 2.21 | 8.0e-31 | positive |
| $\geq 5$ | 277 | 2231 | 1527 | 27324 | 2.22 | 8.0e-27 | positive |
| $\geq 10$ | 191 | 2317 | 827 | 28024 | 2.79 | 3.3e-29 | positive |
| $\geq 20$ | 118 | 2390 | 458 | 28393 | 3.06 | 1.3e-21 | positive |
| $\geq 50$ | 74 | 2434 | 197 | 28654 | 4.42 | 1.9e-21 | positive |
| $\geq 100$ | 53 | 2455 | 87 | 28764 | 7.14 | 6.1e-23 | positive |
| $\geq 300$ | 22 | 2486 | 26 | 28825 | 9.81 | 2.4e-12 | positive |

The dose-response is the cleanest signal in the analysis (Figure 4). Starting from the first comment, the more a benefit is endorsed, the more likely it is backed by trial evidence: while only ∼9% of all endorsed benefits are RCT-demonstrated, ∼38% of heavily-endorsed benefits (≥100 comments) are roughly a four-in-ten chance of RCT backing at the top versus about one-in-nine overall. Strong community consensus is therefore a genuinely useful prior for clinical efficacy.

**Figure 4.**
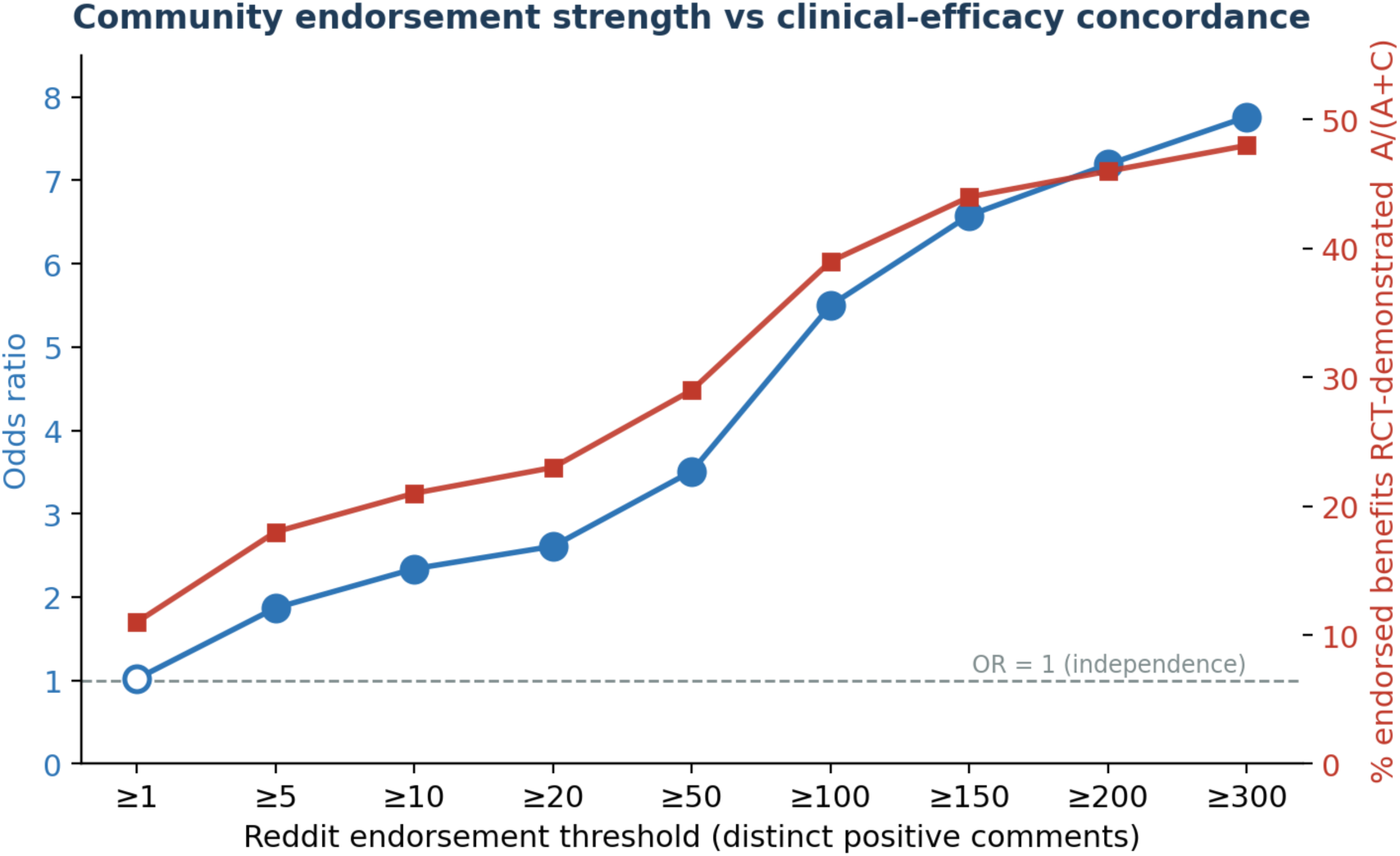
Odds ratio (blue, left axis) and the share of endorsed benefits that are RCT-demonstrated (red, right axis) against the Reddit endorsement threshold. The ≥1 point is open (not significant); all higher thresholds are significant (Fisher p < 0.05).

#### 3.2.2 Sensitivity to the clinical-support definition

At the single-comment threshold every definition is negative or null, because the universe contains many demonstrated benefits for clinical indications users never discuss (inflating cell B). Two successive corrections move the cutoff-1 odds ratio from negative to a true null: crediting published PubMed RCTs removes results-posting bias (0.70 → 0.77), and the abstract-grounded relevance pass removes spurious cross-condition matches that had inflated cell A, lifting the OR to 1.02 (Table 3). The looser “completed” and “any-indication” definitions are strongly negative because they inflate cell B with obscure indication matches and are the least trustworthy.

**Table 3.** Sensitivity to the clinical-support definition at Reddit cutoff ≥1, ordered most to least credible. The recommended primary definition (top row) is direction-aware, posting-bias-free and counts a study only for the indication its abstract shows it tested.

| Clinical-support definition (cutoff $\geq 1$) | A | B | C | OR | $\chi^2$ p |
| --- | --- | --- | --- | --- | --- |
| Efficacy, de-biased + cleaned (PRIMARY) | 802 | 2,540 | 6,605 | 1.02 | 0.60 (n.s.) |
| Efficacy, de-biased, pre-cleaning | 1,538 | 6,092 | 5,869 | 0.77 | $3.1 \times 10^{-16}$ |
| Efficacy, CT.gov posted-results only | 179 | 816 | 7,228 | 0.70 | $2.5 \times 10^{-5}$ |
| Published results (any direction) | 665 | 3,446 | 6,742 | 0.59 | $2.3 \times 10^{-33}$ |
| $\geq 1$ completed trial | 2,694 | 15,660 | 4,713 | 0.30 | $\approx 0$ |
| $\geq 1$ matched trial (any indication) | 3,026 | 19,666 | 4,381 | 0.15 | $\approx 0$ |

#### 3.2.3 Per-ingredient analysis and multiple testing

Of 329 ingredients, 182 had non-degenerate margins and were testable; the remainder had too few claims in one or more cells. Twenty ingredients were nominally significant (p < 0.05), but after Bonferroni correction (α = 2.7×10⁻⁴) only one remained significant. This is the expected consequence of small per-ingredient counts and confirms that the pooled all-pairs test - a single hypothesis requiring no multiple-testing correction - is the appropriate primary analysis.

Illustrative concordant (cell A) pairs include melatonin→sleep-cycle regulation/sleep quality, ginger→nausea relief, niacinamide and zinc→acne support, and L-theanine→anxiety relief; the last correctly recovered only after crediting PubMed RCTs that CT.gov posting bias had hidden. High-volume Reddit-only (cell C) pairs such as vitamin C→acne/collagen and glycine→improved sleep are the priority *research-gap or novel-signal* candidates, alongside vocabulary/paraphrase artifacts (e.g. “melatonin production support”) that should be screened before interpretation.

#### 3.2.4. Identification of the most discussed and clinically validated nutraceutical-health claim pairs

In order to delineate further the Reddit landscape regarding the trends of discussion on nutraceuticals we are presenting the most discussed and clinically validated nutraceutical-health claim pairs (Table 4). We observed in the first positions the melatonin-sleep pairs in different versions. Melatonin is a naturally occurring hormone that regulates circadian rhythm, sleep onset and quality. Melatonin supplements are synthetical but identical to the natural molecule and different clinical studies have proven their efficacy in regulating sleep disturbances.

**Table 4.** The top20 of the most discussed pairs ingredient-claim supported by clinical studies confirmation.

| Rank | Ingredient | Benefit | Reddit_positive_comments | Clinical_support |
| --- | --- | --- | --- | --- |
| 1 | Melatonin | Sleep cycle regulation | 9173 | PubMed |
| 2 | Melatonin | Improved sleep quality | 5693 | CT.gov+PubMed |
| 3 | Ginger | Nausea relief | 2301 | CT.gov+PubMed |
| 4 | Zinc | Acne support | 1150 | PubMed |
| 5 | Niacinamide | Acne support | 1150 | PubMed |
| 6 | L-theanine | Anxiety relief | 1053 | PubMed |
| 7 | Melatonin | Sleep health | 818 | CT.gov+PubMed |
| 8 | L-theanine | Improved sleep quality | 668 | PubMed |
| 9 | L-theanine | Sleep cycle regulation | 652 | PubMed |
| 10 | Ashwagandha | Anxiety relief | 582 | PubMed |
| 11 | L-theanine | Relaxation/calming effects | 569 | PubMed |
| 12 | Melatonin | Insomnia | 481 | CT.gov+PubMed |
| 13 | Zinc | Acne | 450 | PubMed |
| 14 | Curcumin | Anti-inflammatory effects | 427 | PubMed |
| 15 | GABA | Anxiety relief | 377 | PubMed |
| 16 | Maca | Libido enhancement | 359 | CT.gov |
| 17 | Ashwagandha | Improved sleep quality | 358 | PubMed |
| 18 | Zinc | Diaper rash | 344 | PubMed |
| 19 | Curcumin | Reduced inflammation | 339 | PubMed |
| 20 | GABA | Improved sleep quality | 333 | CT.gov+PubMed |

#### 3.2.5 User variability of response on supplements

The concordance analysis so far counts a claim as “endorsed” when enough users report a benefit, but it ignores whether users *agree*. For some claims almost everyone reports a benefit; for others, reports are split or mostly negative. We capture this with an agreement score for each (ingredient, benefit) pair:

*consensus = positive reports / (positive + no-effect reports),*

calculated over the 711 pairs with at least 20 directional comments. A consensus near 1 means users overwhelmingly report a benefit; near 0 means most report no effect. (“No-effect” means the benefit was not obtained)

Agreement differs far more between claims than chance alone would produce. Grouping the 711 pairs by whether their 95% confidence interval lies above, across, or below 0.5 as represented in Table 5.

**Table 5.** Distribution of the 711 claim pairs grouped by level of agreement with the Reddit community reports.

| Group | Claims | Share |
| --- | --- | --- |
| Consensus-positive (users mostly agree it helps) | 573 | 81% |
| Contested (reports split near 50/50) | 55 | 8% |
| Consensus-negative (users mostly report no effect) | 83 | 12% |

Most claims are consensus-positive, but the distribution is clearly three-peaked (Fig. 5): a large high-agreement mode (437 claims at consensus ≥ 0.90), a small contested band near 0.5, and a low cluster of refuted claims (73 claims below 0.30).

**Figure 5.**
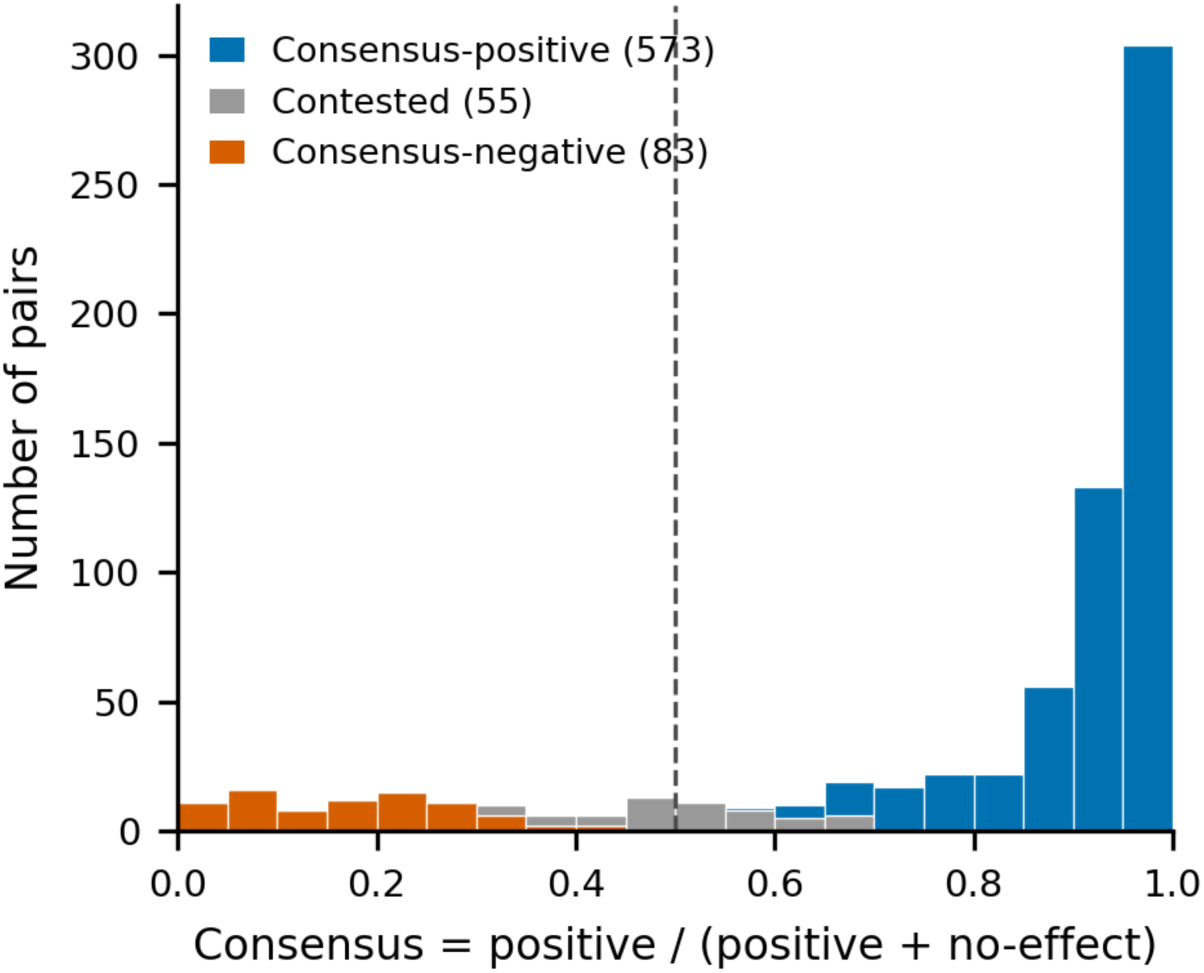
Agreement (consensus) across the 711 claims, with bars coloured by group. Agreement is high for most claims, but a contested band near 0.5 and a refuted cluster below 0.3 stand clearly apart.

The groups make biological sense. High-agreement claims are mostly well-known, single-purpose uses (melatonin→sleep, ginger→nausea). Contested claims (≈0.5) are dominated by vague or paraphrased benefit terms (“serotonin support”) and by caffeine, whose effects genuinely differ between people. Refuted claims are ones users actively contradict, caffeine making anxiety or reflux *worse*, or rejected skin/anti-aging claims (Table 6). The refuted group is the most trustworthy signal: because users are more likely to post when something works, a positive rate as low as 16/687 (caffeine causing anxiety) is hard to explain as anything but genuine contradiction. Contestation is therefore a useful flag for claims that are ill-posed or disputed, which can be screened out before further use.

**Table 6.** Representative nutraceuticals-health claim pairs for each of the group of Reddit community agreement.

| Group | Representative claims (positive / total reports) |
| --- | --- |
| **Near-unanimous positive** | Berberine→insulin sensitivity (99/99)<br>Zinc→skin health (98/98)<br>Lavender→calming (98/98)<br>Ginger→nausea (2,301/2,343)<br>Quercetin→anti-inflammatory (79/79) |
| **Contested ($\approx 0.5$)** | Melatonin→“melatonin production support” (0.51)<br>Caffeine→appetite/digestion (0.49)<br>Caffeine→migraine (0.48)<br>L-tyrosine→serotonin support (0.48) |
| **Consensus-negative (users refute)** | Pea-peptides→anti-aging (1/100)<br>Tart cherry→hyperuricemia (0/20) |

Agreement carries information beyond a simple endorsement count. Claims with clinical support show modestly higher agreement than those without (median 0.95 vs 0.92; Fig. 6). This difference remains after accounting for how much each ingredient is discussed overall, so it is not merely that famous, heavily-discussed ingredients are both agreed-upon and well-studied. The effect is real but small - the two groups overlap substantially - so agreement is best used to help *rank* claims for follow-up, not to predict clinical success on its own.

**Figure 6.**
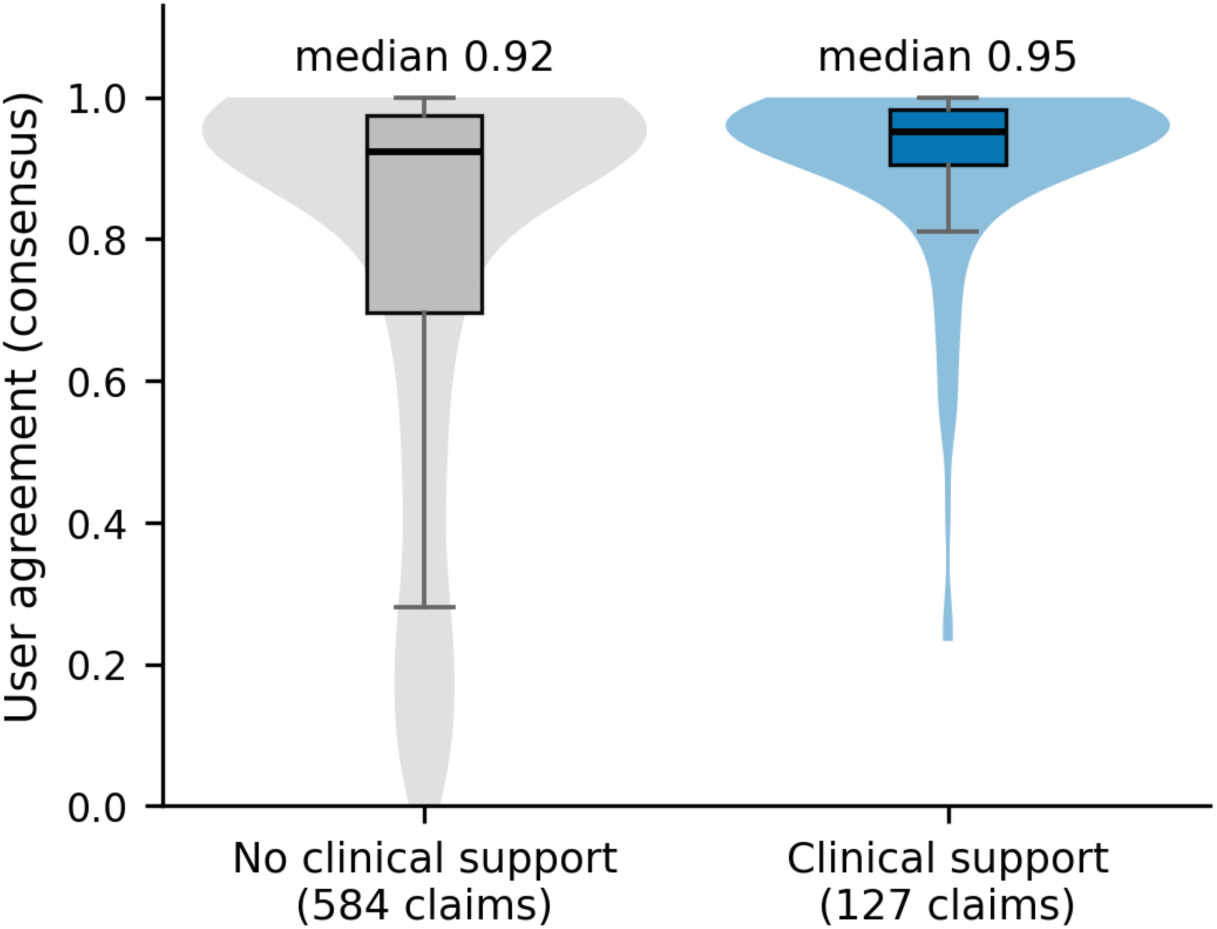
User agreement for claims with versus without clinical support (a significant trial or RCT result for that exact indication). Supported claims have modestly higher agreement, but the distributions overlap heavily, so the score ranks rather than predicts.

The heterogeneity and contestation we observe in this analysis are consistent with, and help prioritize, the personalization of nutraceuticals, a hypothesis grounded in established baseline- and genotype-dependent response (following the Vitamin D literature on benefiting the baseline deficient people or caffeine’s connection with CYP1A2/ADORA2A responder phenotypes)

## 4. Discussion

This study provides, to the best of our knowledge, the first systematic framework for evaluating social-media-derived RWE in the nutraceutical domain by directly linking community-reported benefits to the clinical-trial evidence base on a shared benefit vocabulary. The central finding is a threshold-dependent relationship: clinical efficacy and community endorsement are independent at the single-comment noise floor but positively concordant once a benefit is endorsed by at least two distinct users, with concordance strengthening monotonically as consensus grows. The practical implication is that the strength of community consensus, not its mere presence, is the informative quantity. A benefit endorsed by a hundred or more users carries roughly a four-in-ten prior probability of RCT-demonstrated efficacy, several-fold above baseline.

Two methodological points were decisive and generalize to other social-media RWE work. First, results-posting bias was the dominant artifact: because only ∼15% of trials post structured results to CT.gov, and posting is skewed toward industry/regulatory trials, heavily-studied but rarely-posted supplements were initially mis-scored as unsupported, manufacturing an apparent negative association. Crediting the published literature via PubMed corrected this. Second, over-broad condition crosswalks produced spurious cross-condition matches that inflated apparent concordance; an abstract-grounded relevance pass was needed to count a study only for the indication it actually tested. Analyses that rely on automated condition mapping without reading the underlying evidence risk both biases.

The observational mismatch between discussion and research is itself a finding. The community concentrates on sleep, skin and anxiety while the trial literature concentrates on cardiovascular, pain, metabolic and mood endpoints. High-volume, clinically-unsupported (cell C) pairs are the natural output of this framework: a prioritized, consensus-weighted list of hypotheses, either genuine research gaps deserving formal study or vocabulary artifacts to be screened out.

Extant research and our results position social media as offering direct access to patient experience at a scale and immediacy that conventional surveys and spontaneous-reporting systems cannot match. However, establishing causality, or validating any finding that suggests it, remains challenging. Self-selection, the absence of denominators and verified exposure, paraphrase and normalization noise, and the lack of controlled confounders mean these signals are hypothesis-generating rather than confirmatory. The appropriate role is a complementary, front-end prioritization layer that feeds, rather than replaces, formal experimental research.

Interestingly, Reddit community-reported experience are in high agreement with the outcome of the clinical trials. The highest level of concordance is observed in well-known natural ingredients-health claim pairs such as zinc-skin, likely due to its anti-inflammatory properties, lavender-calming effect, etc. On the other hand, there are nutraceuticals-health claim pairs that are under debate in Reddit community for their correlation, such as caffeine-migraine and pea-peptides-anti-aging effect. This set of observations points towards the need for personalization in the nutraceutical field.

## 5. Conclusion

Across 329 ingredients and 31,359 (ingredient, benefit) pairs, clinically demonstrated benefits and Reddit endorsement are independent at the single-comment noise floor but positively associated once a benefit is endorsed by multiple users, with concordance rising monotonically with consensus (odds ratio ∼7.1 at ≥100 endorsing comments). Social-media forums such as Reddit are therefore a valuable, consensus-weighted source of real-world data for the nutraceutical domain; best used as a complementary, hypothesis-generating prior that prioritizes ingredients, indications and doses for formal study, rather than as a substitute for controlled experimental evidence.

## Data Availability

All data produced in the present work are contained in the manuscript

